# An outbreak inside an outbreak: rising incidence of carbapenem-resistant isolates during the COVID-19 pandemic. Report from a tertiary care center in Argentina

**DOI:** 10.1101/2021.11.11.21266237

**Authors:** Maximiliano Gabriel Castro, Lucía Ubiergo, Macarena Vicino, Gisel Cuevas, Fernanda Argarañá

## Abstract

**Introduction:** COVID-19 outbreaks have left us to deal with an aftermath on many fronts. In particular, disproportionate use of antibiotics, high ICU burden and longer in-hospital stays during the pandemic have been proposed to aggravate the emergency posed by carbapenem-resistant isolates (CRI), specially through carbapenemase production. However, there have been few reports worldwide regarding changes in CRI incidence and little latinamerican literature.

**Objective:** We set out to determine whether the incidence of CRI rose in a tertiary care center in Santa Fe, Argentina during the time period with active cases of COVID-19.

**Methods:** Analytic epidemiologic study retrospectively designed. Two time periods were defined: P1 (without active cases of COVID-19) from September, 2019 to August, 2020 and P2 (starting at the onset of the first wave of COVID-19 in this institution) from September, 2020 to June 2021. All clinically-relevant microbiological samples -those meant for diagnostic purposes-taken during the study period from patients in the Internal Medicine and Surgical wards as well as the Intensive Care Units were included. Incidence was calculated by dividing the number of CRI during each time frame by the count of patient-day during that same period, multiplied by a hundred.

**Results:** 9,135 hospitalizations, 50,145 patient-days of analysis. A total of 7285 clinical samples were taken, with an overall positivity for CRI of 12.1% (n=883). Overall CRI incidence during P2 was 2.5 times higher than in P1 (2.52 vs 0.955/100 patient-days, p <0.001). ICU CRI incidence raised from 6.78 to 8.69/100 patient-days in P2 (p=0.006).

**Conclusion:** We found alarming rates of CRI in our center, 2.5 times higher than previous to the first COVID-19 wave, similar to other reports worldwide. To our knowledge, this is one of the few Latin-American reports on the effect of the COVID-19 pandemic on CRI incidence.

## INTRODUCTION

In november 2019, a cluster of patients with pneumonia caused by the severe acute respiratory syndrome coronavirus 2 (SARS-CoV-2) was reported in Wuhan, province of Hubei, in the Republic of China. Now that widespread vaccination has helped to control the pandemic, we are facing the aftermath on many fronts. Regarding infectious diseases, we are yet to assess what landscape of antibiotic resistance we will have to face.

In cases of seasonal influenza, as well as during pandemic outbreaks, development of concomitant bacterial lung infections was a common finding, predominantly in severe cases. This was shown in a historical review of pathological reports of 1918-1919 influenza outbreaks(1) and by a CDC report of autopsy findings during the H1N1 pandemic(2)-that were later reinforced by a systematic review (3).

In contrast, previous studies regarding bacterial infections in COVID-19 are scarce. Moreover, they seldom specify any critical information, including diagnostic criteria, timing of sample collection and the identity of bacterial species (4). Two meta-analyses showed an incidence of bacterial infections of between 7-8% in hospitalized COVID-19 patients, which may escalate up to 16% in critically-ill patients(5–7). However, estimates of antibiotic administration reached approximately 70%(8,9).

There are multiple examples of this disproportionate use of antibiotics (10–12), that ranges from as low as 33.7% to around 90% in hospital settings with predominance of broad-spectrum antibiotics (13,14), even in facilities with strong Antibiotic Stewardship Programs. Nevertheless, from nationwide perspectives, this may have subsided in the outpatient setting due to an overall trend in reduction of antibiotic prescription. However data is not available for most countries, including Argentina (15–17).

Particularly in England, even with a marked decline in overall antibiotic consumption, the amount of prescribed doses of antibiotics per hospital admission increased during the COVID-19 pandemic(18). This may not only be in line with early empiric antibiotic treatment but also with the fact that COVID-19 patients have been described to have longer Intensive Care Unit (ICU) stays, with longer need for mechanical ventilation and more frequent tracheostomies(19), in close relationship to a higher rate of hospital-acquired infections(20) -predominantly ventilator-associated pneumonias(21,22).

Greater use of antibiotics in hospital settings, which are the niche for multidrug-resistance development, may add to the already complex situation regarding carbapenem resistance, which appears to be rising (23,24), and which is mainly associated with antibiotic misuse. In particular, resistance to imipenem among *Klebsiella pneumoniae* isolates in Argentina has grown in the last decade, reaching 19.9%(25).

On top of everything, the need for new personnel and the redistribution of the existing staff to cope with the pandemic, along with the spread of certain practices such as the use of double pair of gloves -out of fear of contagion at the start of the pandemic- and the lack of resources for Antibiotic Stewardship Programs may have pressured antibiotic resistance. However, there is an increasing albeit scarce evidence available on this matter, specially in Latin-America and the Caribbean. Worldwide, some reports have appeared highlighting the threat of multidrug-resistant pathogens during the COVID-19 pandemic(26).

We therefore set out to determine whether the incidence of carbapenem-resistant isolates (CRI) rose in a tertiary care center in Argentina during the time period with active cases of COVID-19.

## METHODS

This is an analytic epidemiologic study retrospectively designed in order to assess the incidence of CRI in microbiological samples of clinical relevance, in a tertiary-care center from Santa Fe, Argentina during the COVID-19 pandemic, compared to a previous time period.

Two time periods were defined: P1 (without active cases of COVID-19) from September, 2019 to August, 2020 and P2 (starting at the onset of the first wave of COVID-19 in this institution) from September, 2020 to June 2021.

All clinically-relevant microbiological samples -those meant for diagnostic purposes-taken during the study period from patients in the Internal Medicine and Surgical wards as well as the Intensive Care Units were included. Therefore, rectal swabs as well as all surveillance samples were excluded. Sample collection was decided by the treating physician and the interpretation of these results was not investigated due to the volume of samples.

Blood samples from peripheral venipunctures and those drawn from catheter lumens were put into Bact/ALERT^®^ (bioMérieux, Argentina) blood culture aerobic bottles. A blood culture set, for the purpose of this report is considered to be composed of one to three blood culture aerobic bottles taken at once. Blood samples taken from catheter lumens were treated similarly.

Urine samples, catheter tips and respiratory samples (either sputum or bronchoalveolar lavage fluids) were also included.

Various fluids culture (VFC) is composed of clinical samples extracted from synovial fluid, ascitis, pleural fluid or pericardial fluid, while various materials culture (VMC) is composed of clinical samples extracted from abscesses and diverse tissues.

All clinical samples were processed and analyzed according to international standards.

The presence of bacterial growth from blood culture bottles was done through Bact/ALERT 3D System (bioMérieux, Argentina) with maximum incubation ranging from seven days to 14 days, in ICU specimens or in those with suspected slow-growth microorganisms. Positive blood culture bottles were afterwards sub-cultured in agar plates.

The rest of clinical samples were grown in standard agar plates, and positivity was defined according to standard CFU cut-off value.

Bacterial identification and susceptibility testing were performed with the automated system Vitek 2C (bioMérieux, Argentina). Resistance to carbapenems was defined according to cutoff values on CLSI M100-ED31:2021 Performance Standards for Antimicrobial Susceptibility Testing.

The primary analysis of interest was the differential incidence between the two time periods, overall and in the ICU wards in particular.

Incidence was calculated by dividing the number of CRI during each time frame by the count of patient-day during that same period, multiplied by a hundred. Patient-day was the sum of the in-hospital length of stay -measured in days-of individual patients during a time frame. 95% confidence intervals were calculated using the OpenEpi software.

Comparisons between incidences were performed using Fisher’s exact test. The limit for statistical significance was a two-sided p <0.05.

Statistical analyses were performed both with OpenEpi and SPSS Statistics v27.0. Graphs were generated using Microsoft Excel 365.

## RESULTS

Taken together, both periods included 9,135 hospitalizations, and 50,145 patient-days of analysis. P1 represented 62.8% of hospitalizations but only 48.7% of patient-days, with a monthly decline in hospitalizations in P2 (340 vs 478). The surgical ward, which had the leading number of admissions (60.0%), showed a decline in monthly admissions in P2 (158.2 vs 325). While the Internal Medicine ward showed no significant change in admissions between the periods, three new ICU wards had to be opened (totalling 6 UCI wards) during the P2 due to an increase in monthly admissions (72.6 vs 40.0). ICU mortality rose from 31.7% to 43.3%, and so did mean in-hospital stay, from 4.9 to 8.7 days. Moreover, these estimates may be biased downwards due to an increase in referral to other centers in P2 from 3.76% to 21.8%, since due to scarcity of beds in ICU wards, patients mainly remained in this center during the COVID-19 isolation period.

A total of 7285 clinical samples were taken, the majority of which were blood culture sets (n=3238, with a monthly estimate that raised 1.7 times in P2 at the expense of ICU wards). In second place, clinical samples from respiratory samples and catheter tip also increased by 1.9 and 2.18 times, respectively (Table 1).

**Table 1.** Number of culture samples and positivity rate for CRI, divided by culture type and period. **Bold letters:** significant changes in positivity rates.

Annex
| Type of culture | Period 1 |  | Period 2 |  | Total |  |
| --- | --- | --- | --- | --- | --- | --- |
|  | n | %CRI | n | %CRI | n | %CRI |
| Blood cultures | 1340 sets | **3.58%** | 1898 sets | **9.16%** | 3238 sets | 6.86% |
| Blood cultures from catheter lumen | 147 sets | 10.9% | 216 sets | 11.5% | 363 sets | 11.3% |
| Catheter tip | 214 | **14.0%** | 469 | **21.3%** | 683 | 19.0% |
| Urine culture | 443 | **4.74%** | 504 | **14.4%** | 947 | 9.93% |
| VFC | 360 | 3.33% | 289 | 3.1% | 649 | 3.24% |
| VMC | 203 | **11.3%** | 197 | **4.50%** | 400 | 8.00% |
| Respiratory cultures | 283 | **28.9%** | 545 | **47.7%** | 828 | 41.3% |
| Fecal culture | 96 | 1.04% | 81 | 0.00% | 177 | 0.565% |
| Overall | 3086 | 7.55% | 4199 | 15.2% | 7285 | 12.1% |

A total of 883 CRI were isolated (80.0% from the ICU wards) from 359 patients, which barely tripled during P2 (640 vs 233). Overall CRI incidence was 1.66/100 patient-days (CI 95% 1.55-1.78). Overall CRI incidence during P2 was 2.5 times higher than in P1 (2.52 vs 0.955/100 patient-days, p <0.001). ICU CRI incidence raised from 6.78 to 8.69/100 patient-days in P2 (p=0.006) (Graph 1).

**Graph 1.**
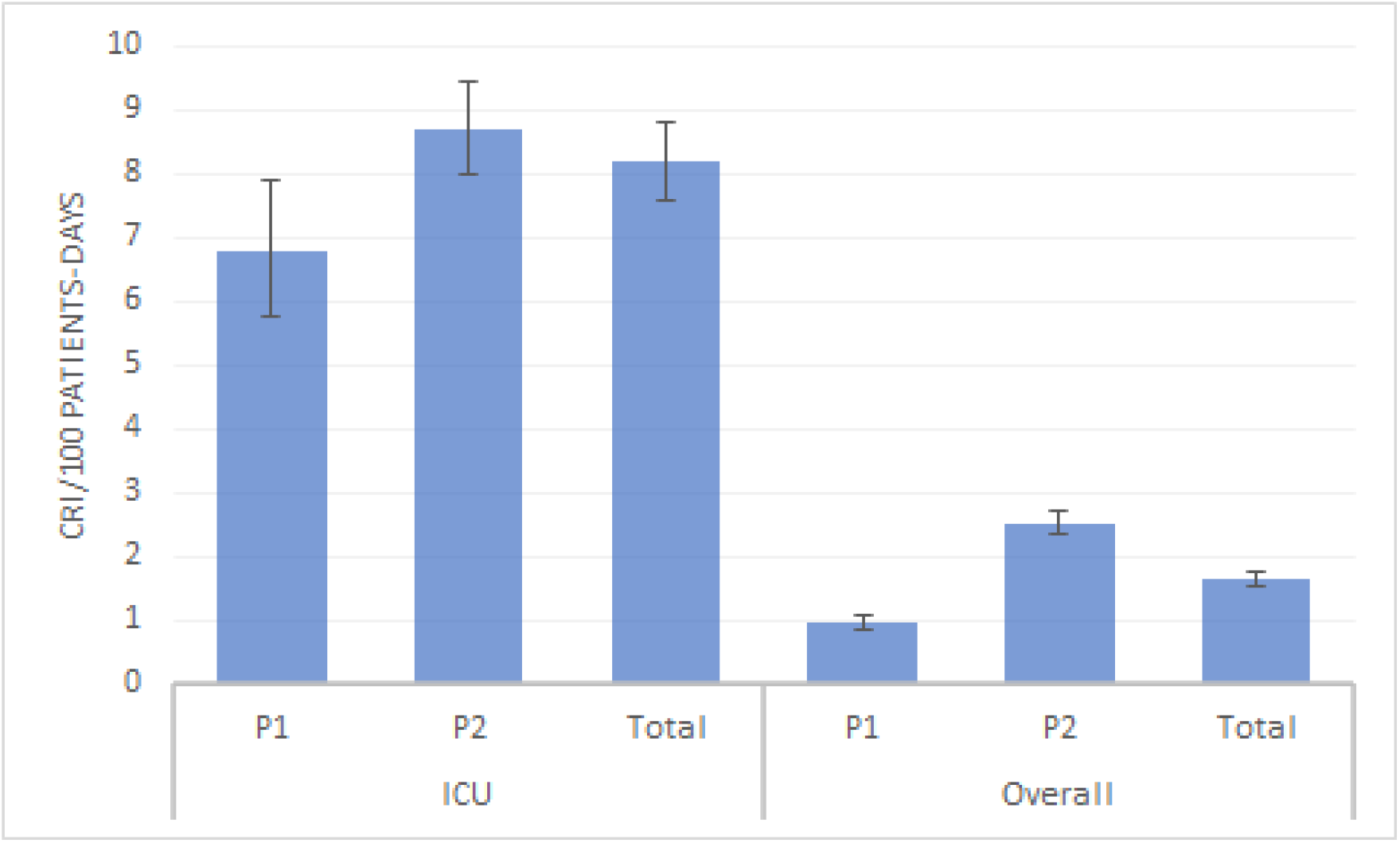
Comparison of incidences (95% CI) of CRI between periods in the ICU wards and overall.

This was due not only to an increment in the number of clinical samples collected, but also due to higher positivity. Overall blood cultures positivity to CRI raised from 3.58% to 9.16%, urine culture positivity from 4.7% to 14.4% and respiratory samples culture from 28.9% to 47.7%. Other changes in positivity rates are shown in Table 1.

The relative frequencies of the bacteria isolated are shown in Table 2.

**Table 2.** Absolute and relative frequencies of isolated microorganisms with carbapenem resistance.

| Microorganism | n | % |
| --- | --- | --- |
| *Klebsiella pneumoniae* | 307 | 36.8 |
| *Acinetobacter baumannii* | 157 | 17.8 |
| *Pseudomonas aeruginosa* | 156 | 17.7 |
| *Proteus mirabilis* | 129 | 14.6 |
| *Serratia marcescens* | 66 | 7.47 |
| *Enterobacter cloacae* complex | 29 | 3.28 |
| *Escherichia coli* | 8 | 0.906 |
| *Pseudomonas putida* | 7 | 0.793 |
| *Morganella morganii* | 6 | 0.680 |
| *Aeromonas hydrophila* | 5 | 0.566 |
| *Proteus penneri* | 3 | 0.400 |
| *Pseudomonas fluorescens* | 3 | 0.400 |
| *Proteus vulgaris* | 2 | 0.227 |
| *Providencia stuartii* | 2 | 0.227 |
| *Aeromonas sobria* | 1 | 0.113 |
| *Chryseobacterium indologenes* | 1 | 0.113 |
| *Pseudomonas aeruginosa mucosa* | 1 | 0.113 |
| Total | 883 | 100 |

Each patient with at least one CRI had a mean of 2.32 isolates during the in-hospital stay. The temporal trends in monthly incidence are shown in Graph 2.

**Graph 2.**
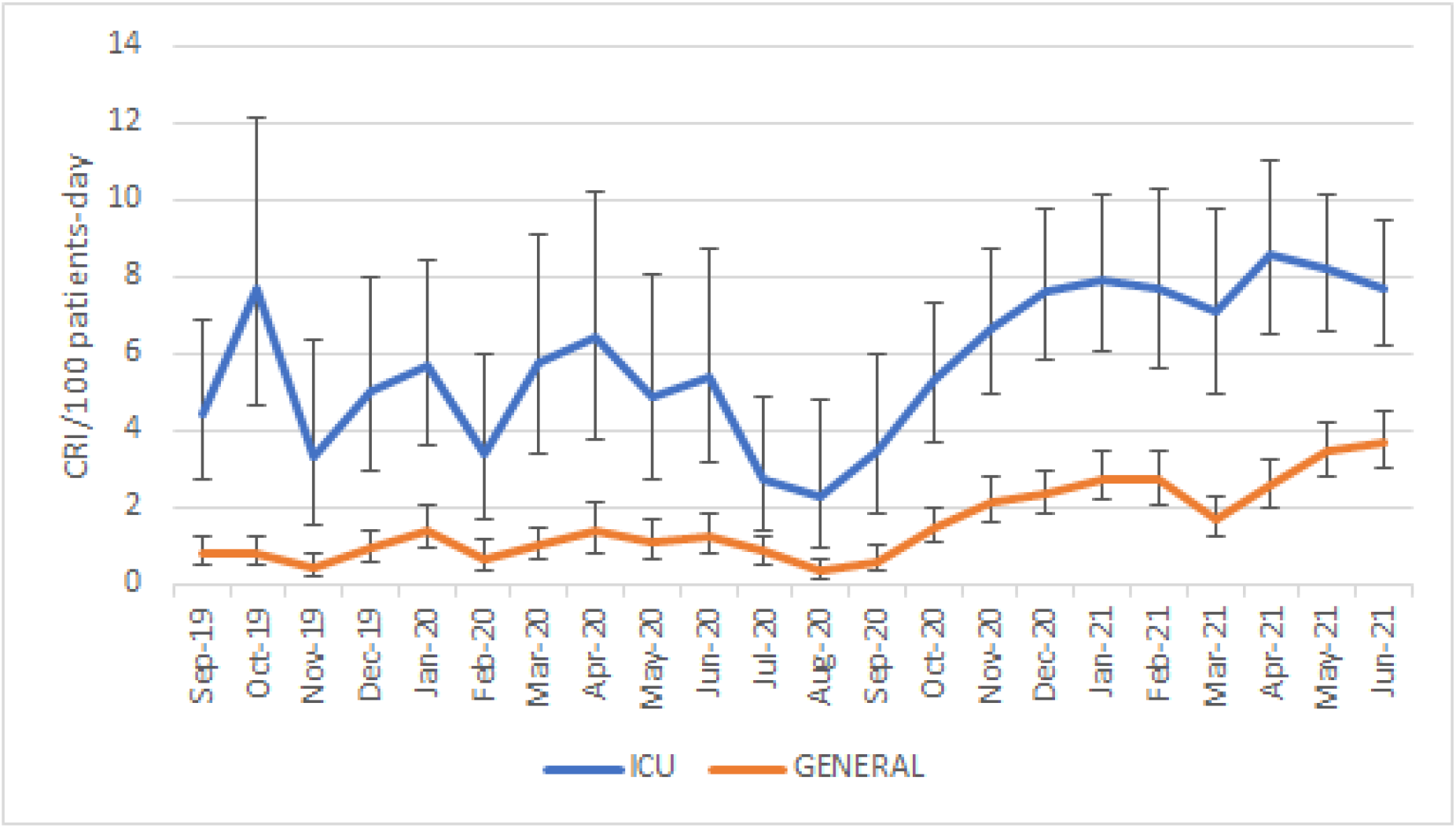
Temporal trends in monthly incidences (95% CI) of CRI in ICU wards and overall.

## DISCUSSION

In this study we hypothesized that during the COVID-19 pandemic there would be an increase in carbapenem resistance incidence in our center, based on the disproportionate use of antibiotics, longer in-hospital stay and the higher ICU occupancy during this period, as well as the worldwide trend in carbapenem resistance growth.

Indeed, we found a 2.5 times increase in CRI incidence overall, at the expense of ICU patients, who not only had longer in-hospital stays but also doubled in number in P2. Bed occupancy has been linked to multidrug-resistant pathogens epidemic(27).

Moreover, each patient with at least one carbapenem resistant microorganism isolated had a mean of 2.3 isolates. Therefore, the fact that a large proportion of critical patients were referred to other centers before ICU discharge may have downsized the incidence of CRI, as opposed to what may have happened had they stayed.

Patient referral may have posed an epidemiologic threat due to dispersion of multidrug resistant microorganisms between facilities that could aggravate the fact that in Santa Fe, Argentina, resistance to imipenem in *Klebsiella sp*. is already 10 points above the national estimate. However, the effect of this phenomenon has not been studied in this context. An epidemiologic study in France previous to the pandemic showed that patient transfer between facilities sustained multidrug-resistant pathogens epidemics(28).

The most common CRI was *Klebsiella pneumoniae* -as described in other reports(29), followed by *Acinetobacter baumanii, Pseudomonas aeruginosa* and *Proteus mirabilis*. Carbapenem-resistant *Acinetobacter baumanii*, in particular, is a specially dangerous threat to ICU patients and there have already been reports of outbreaks during the COVID-19 pandemic(30).

Even in the absence of official Latin-American data on carbapenem-resistance during the COVID-19 pandemic, many centers worldwide have reported a surge in CRI, albeit with heterogeneous methodologies and only a few of them with longitudinal data. An Italian report from 2020 showed a surge in CRI incidence, even with a strong infectious control program and an overall decreasing trend from previous years(31). Similar to this, a New York city center showed a tendency to a greater incidence of CRI in COVID-19 patients, after a steady decline in previous years (32). The majority of isolates in this report were from respiratory samples, similar to what happened in our center.

Another Italian prospective observational study published in 2020 reported 109 superinfections in 69 patients (21.9% of hospitalized COVID-19 patients), 44.9% of which were *Enterobacterales*, the majority of which (52/66) were CRI (33). This was in line with an Italian multicentre before-after cross sectional study that found a 7.5 and 5.5-fold increase in incidence rate ratio of colonization and infection, respectively(34).

Regarding Latin-American countries, one prospective cohort study from a tertiary care center in Mexico City that included 794 patients with severe COVID-19, identified 110 hospital-acquired infections in 74 patients, the majority of which (69.6%) were caused by *Enterobacterales*, however with a low prevalence of carbapenem resistance(35).

Recently, the Panamerican Organization of Health published a document warning about the emergence and increment of carbapenemase-producing *Enterobacterales* in Latin-America. There, it highlighted the first report of *Enterobacterales* co-producing KPC and NDM carbapenemases in Argentina -as well as in other countries- and a three times increment in KPC and NDM producing *Enterobacterales* in Uruguay(36).

Several factors may have favoured the rise of CRI incidence in our, as well as other, centers. Limited resources worldwide, as well as high patient burden, may have resulted in difficult decision-making, limiting the use of gloves and gowns to certain situations. Moreover, the use of personal protective equipment created a false sense of security for the personnel tending to cohorted COVID-19 patients. This false sense of security is a particularly dangerous epidemiological threat, since it has already been shown that there is a high likelihood of carriage of carbapenem-resistant *Enterobacterales* in gowns and gloves that come in contact with carriers of CRI (37). On top of that, some have reported, in the context of scarce resources, to have stopped routine rectal screening during the COVID-19 pandemic(38), therefore not being able to identify the patients that pose the highest risk, Even more, during the COVID-19 pandemic, contact and respiratory isolation for all patients posed a significant burden on the health personnel. A high percentage of patients in contact isolation has shown to reduce compliance to hand hygiene(39). Overcrowding, under-staffing and a high patient-caregiver ratio -all common during outbreaks of COVID-19 pneumonia-have been associated with a higher risk of cross-transmission.(40)

## CONCLUSION

In the midst of the COVID-19 pandemic, several factors were thought to favour outbreaks of carbapenem resistance, which had already been declared a worldwide emergency. Indeed, we found alarming rates of CRI in our center, 2.5 times higher than previous to the first COVID-19 wave, similar to other reports worldwide.

To our knowledge,to this date this is one of the few Latin-American reports on the effect of the COVID-19 pandemic on CRI incidence. More studies are needed to understand the real trend in carbapenem resistance and whether unified efforts in infectious control measures will be able to manage these outbreaks in the aftermath of this pandemic.

## Data Availability

All data produced in the present study are available upon reasonable request to the authors

